# Naturalistic drug cue reactivity in heroin use disorder: orbitofrontal synchronization as a marker of craving and recovery

**DOI:** 10.1101/2023.11.02.23297937

**Authors:** Greg Kronberg, Ahmet O. Ceceli, Yuefeng Huang, Pierre-Olivier Gaudreault, Sarah G. King, Natalie McClain, Nelly Alia-Klein, Rita Z. Goldstein

## Abstract

Movies captivate groups of individuals (the audience), especially if they contain themes of common motivational interest to the group. In drug addiction, a key mechanism is maladaptive motivational salience attribution whereby drug cues outcompete other reinforcers within the same environment or context. We predicted that while watching a drug-themed movie, where cues for drugs and other stimuli share a continuous narrative context, fMRI responses in individuals with heroin use disorder (iHUD) will preferentially synchronize during drug scenes. Results revealed such drug-biased synchronization in the orbitofrontal cortex (OFC), ventromedial and ventrolateral prefrontal cortex, and insula. After 15 weeks of inpatient treatment, there was a significant reduction in this drug-biased shared response in the OFC, which correlated with a concomitant reduction in dynamically-measured craving, suggesting synchronized OFC responses to a drug-themed movie as a neural marker of craving and recovery in iHUD.

## Introduction

Neurobiological theories of drug addiction have proposed that motivational salience is attributed to drug cues at the expense of other reinforcers (e.g., social, food, sex), a key mechanism that perpetuates compulsive drug use and the addiction cycle^1,2^. Supporting these theories, drug cues elicit hyper-reactivity in several brain networks, which correlates with drug craving ratings across substance use disorders (SUD)^3^. These drug cue-reactivity studies typically compare group-averaged [SUD vs. healthy control (HC)] responses to static images sampled from different categories (drug vs. non-drug)^4^. However, the static and decontextualized nature of cues in this paradigm makes it difficult to capture three key aspects of the imbalanced salience attribution as it manifests in the real world: 1) richness and complexity of real environments: several important components of naturalistic drug use environments are difficult to simulate with static images alone, including drug seeking/purchasing, preparation, use, and the ensuing high (and the integration of these components over time); 2) direct competition for salience attribution between drug and non-drug cues: during natural experience these cues are embedded within the same dynamic environmental context; and 3) the social nature of addiction and recovery: peers can influence each other through shared interpretation of their environment, which can be reflected in shared brain responses^5^. Image-based cue reactivity studies typically average responses over many trials however, precluding the measurement of shared responses, and missing this important social dimension^5^. Therefore, despite several decades of research, we still cannot answer whether complex drug cues would outcompete other stimuli if both types of cues are directly compared within the same dynamic context, or describe how brain responses are shared within groups, as would occur in real-world interactions.

Movies establish a contiguous complex narrative context and maximize engagement, driving attentional, motivational, and higher order cognitive processes^6^. In particular, a drug-themed movie could facilitate a deeper engagement with a simulated drug environment in people with SUD, requiring parallel processing of drug and non-drug scenes embedded in the same narrative, which would more closely capture their mutual competition for salience attribution in the real world. Moreover, analyses of movie fMRI data allow inspection of shared stimulus-locked brain synchronization across individuals, a measure sensitive to complex features of the stimulus and their integration over multiple timescales^7^, and to shared social experience^5^. The latter is especially important in addressing the distinctly social nature of real-world drug use and recovery, allowing a glimpse into the way groups of individuals with common SUD experiences collectively attribute incentive salience in their environment. Despite the promise of such overall improved ecological validity, along with several technical advantages^8^, there have been no studies measuring shared responses to a naturalistic stimulus in any SUD.

Heroin use disorder (HUD) and its neurobiological substrates have been particularly understudied compared to other SUDs^3^. Despite the magnitude of the opioid epidemic, where opioid-related overdose deaths in the United States have increased almost four-fold in the last decade, reaching 80,000 in 2022^9^, fMRI drug cue-reactivity studies in individuals with HUD (iHUD) remain scarce^3^. Moreover, while early evidence suggests the potential for structural and functional neural recovery with treatment across SUDs, there are still very few longitudinal studies assessing the effects of treatment-based abstinence in iHUD (see Parvaz et al. for a recent review)^10^. Specifically, although we recently reported evidence of recovery of inhibitory control prefrontal cortical (PFC) functions in iHUD^11^, there are no longitudinal studies tracking cue reactivity in iHUD, and none that use engaging naturalistic stimuli for the study of this group’s shared brain responses and their potential change with treatment.

Here we measure shared fMRI responses to a heroin-related movie in a sample of inpatients with HUD, studied before and after approximately 3 months of treatment; a sex/age matched sample of HC was scanned at similar time intervals. Specifically, we adapted a reverse correlation method^12^ to identify the movie content that elicited synchronized fMRI responses in each group. We then measured the degree of shared bias towards drug content, when drug and non-drug stimuli are presented within the same dynamic narrative context. We also measured self-reported drug craving, a typical well-validated, easy to measure treatment outcome that changes with abstinence in iHUD^13–15^, predicts future drug use and relapse across SUDs^16^, and correlates with drug cue-reactive fMRI signals predominantly in regions related to reward and salience processing^17^ (although see ^18^). We found that the orbitofrontal cortex (OFC), a region implicated extensively in dysfunctional reward processing and salience attribution, disadvantageous decision making, and craving in addiction, showed synchronized responses that were biased towards drug content in the iHUD. This OFC shared drug-bias was significantly reduced with treatment/abstinence, and changes in OFC synchronization were correlated with changes in craving with treatment. Overall, using a naturalistic and longitudinal design in HUD, we find support for prominent addiction theories on salience attribution, bridging our results with extensive experimental evidence on drug cue-reactivity in other drugs of abuse. Providing much needed data in iHUD, for the first time we highlight group synchronized OFC responses to naturalistic drug cues as a potential neural marker of craving and recovery with treatment in this devastating disorder.

## Results

### Whole brain reverse correlation

Functional MRI blood-oxygen-level-dependent (BOLD) activity was recorded in 30 treatment-seeking medication-stabilized iHUD (40.4±10.3 years, 23 Male, 19 White) and 25 age, sex, and race-matched HC (43.8±10.3 years, 10 Male, 11 White), while subjects watched the first 17 minutes of the movie “Trainspotting”; all subjects returned an average of 15 weeks later (see table 1 for days between sessions) to repeat the same procedures. The overarching narrative structure of the movie is centered around the drug addiction cycle itself, including more abstract elements relevant to HUD (e.g., social, emotional, and economic challenges of addiction). Importantly, the movie clip contains scenes of explicit heroin use, including heroin seeking/purchasing, preparing, using, and feeling high, as well as food, social interactions, and other scenes that are highly salient. To identify stimuli evoking shared responses between subjects within a group, we used an approach similar to Hasson et al.^12^, which was inspired by reverse correlation analyses of single unit recordings in sensory neurons^19,20^. This method first identifies repetition times (TRs) where the BOLD signal is synchronized across individuals, defined here as a significantly non-zero group median signal (tested independently at each TR and corrected for multiple comparisons over TRs). Adjusting for the hemodynamic response, these synchronized TRs are then mapped to 1 second movie bins, which were labeled as drug or non-drug a priori by independent raters (see methods for criteria). To identify the brain regions that exhibited a bias to a certain content in the movie (drug vs. non drug), we then defined a group’s drug bias as the fraction of bins labeled as containing drug (vs. non drug) content for each of 450 regions of interest (ROIs). Drug bias therefore reflects the degree to which group-synchronized BOLD responses follow drug (vs. non drug) content in the movie. This metric was then compared between groups via permutation testing and the resulting p-values were then FDR corrected over all ROIs (Figure 1).

**Table 1.** Sample profile. Significant group differences (corrected for familywise error, α =.05/12=.0041) are flagged with an asterisk. HC: healthy control group; HUD: heroin use disorder group; t: student’s t statistic; χ^2^: chi-square statistic; U: Mann-Whitney test statistic.

|  | HC (n=25) | HUD (n=30) | Group comparison |
| --- | --- | --- | --- |
| Age | 39.97 ± 10.76 | 40.00 ± 8.41 | t(53)=-0.011; p=0.99 |
| Sex (Female/Male) | 9/16; | 6/24; | $\chi^2(1)=1.0$; p=0.31 |
| Race (Black/Other/White) | 9/4/12 | 2/6/22 | Fisher’s exact p=0.027 |
| Education (years) | 16.28 ± 2.89 | 12.24 ± 1.99 | U=650.0; p=5.2e-07* |
| Verbal IQ | 110.28 ± 6.85 | 96.53 ± 11.73 | t(53)=5.2; p=3.7e-06* |
| Nonverbal IQ | 11.96 ± 2.76 | 10.43 ± 3.15 | t(53)=1.9; p=0.064 |
| Beck’s Depression Inventory-II | 3.36 ± 4.69 | 14.30 ± 11.08 | U=110.0; p=6.3e-06* |
| Beck’s Anxiety Inventory | 2.24 ± 4.05 | 10.60 ± 9.47 | U=140.0; p=4.3e-05* |
| Fagerström Test for Nicotine Dependence | 0.04 ± 0.20 | 3.23 ± 2.05 | U=54.0; p=5.4e-09* |
| Years of regular marijuana use | 0.68 ± 1.62 | 10.08 ± 9.09 | U=110.0; p=2.5e-06* |
| Short Michigan Alcohol Screening Test | 0.48 ± 1.12 | 2.60 ± 3.21 | U=150.0; p=5.8e-05* |
| Days between sessions | 98.92 ± 52.90 | 107.20 ± 37.50 | U=270.0; p=0.089 |
| Heroin Craving Questionnaire |  | 38.87 ± 13.50 |  |
| Heroin use (days/past 30) |  | 0.27 ± 0.78 |  |
| Years of regular heroin use |  | 8.48 ± 5.57 |  |
| Heroin Severity of Dependence Scale |  | 10.90 ± 3.79 |  |
| Short Opiate Withdrawal Scale |  | 2.53 ± 2.90 |  |
| Days abstinent from heroin |  | 182.90 ± 199.78 |  |
| Urine toxicology (methadone/suboxone) |  | 26/4 |  |
| Heroin administration (injected/nasal/oral/inhaled) |  | 16/12/1/1 |  |
| Methadone dose (mg) |  | 105.30 ± 54.90 |  |

**Figure 1.**
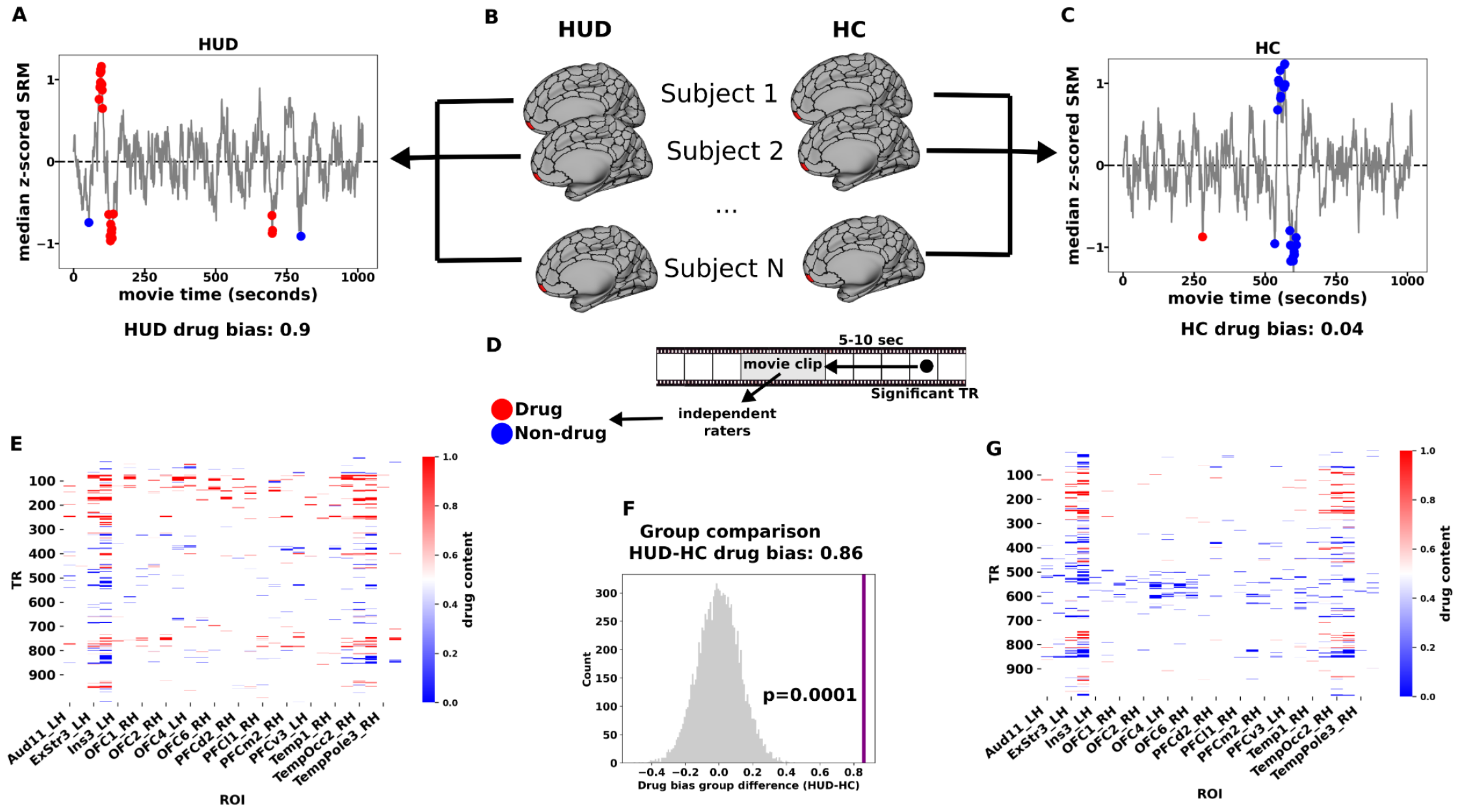
Whole-brain reverse correlation to identify drug-biased processing. An example ROI in the right medial OFC is highlighted to illustrate the method. Within each ROI and group (B), significantly synchronized TRs were identified by fluctuations in the group median signal above a noise threshold (A,C; see Methods Identifying synchronized TRs and region-specific reactivity). Each significant TR was mapped to the 5 second movie clip that preceded that TR by 5-10 seconds. The 5-second clip was broken into1-second bins and each bin was labeled as containing drug-related content (red) or not (blue) (D). Drug bias was then determined as the fraction of significant bins that contained drug content and a test statistic was constructed as the difference in drug bias between the groups (HUD minus HC). This test statistic was compared to a null distribution based on randomly distributing the same number of bins 10,000 times yielding a p-value for each ROI (F), which were then FDR corrected over 450 ROIs. (E,G) Carpet plot of significant TRs in ROIs that showed a significantly greater drug bias in HUD at baseline. Each TR is colored according to the drug content of its associated 5-second clip of the movie.

**Figure 2.**
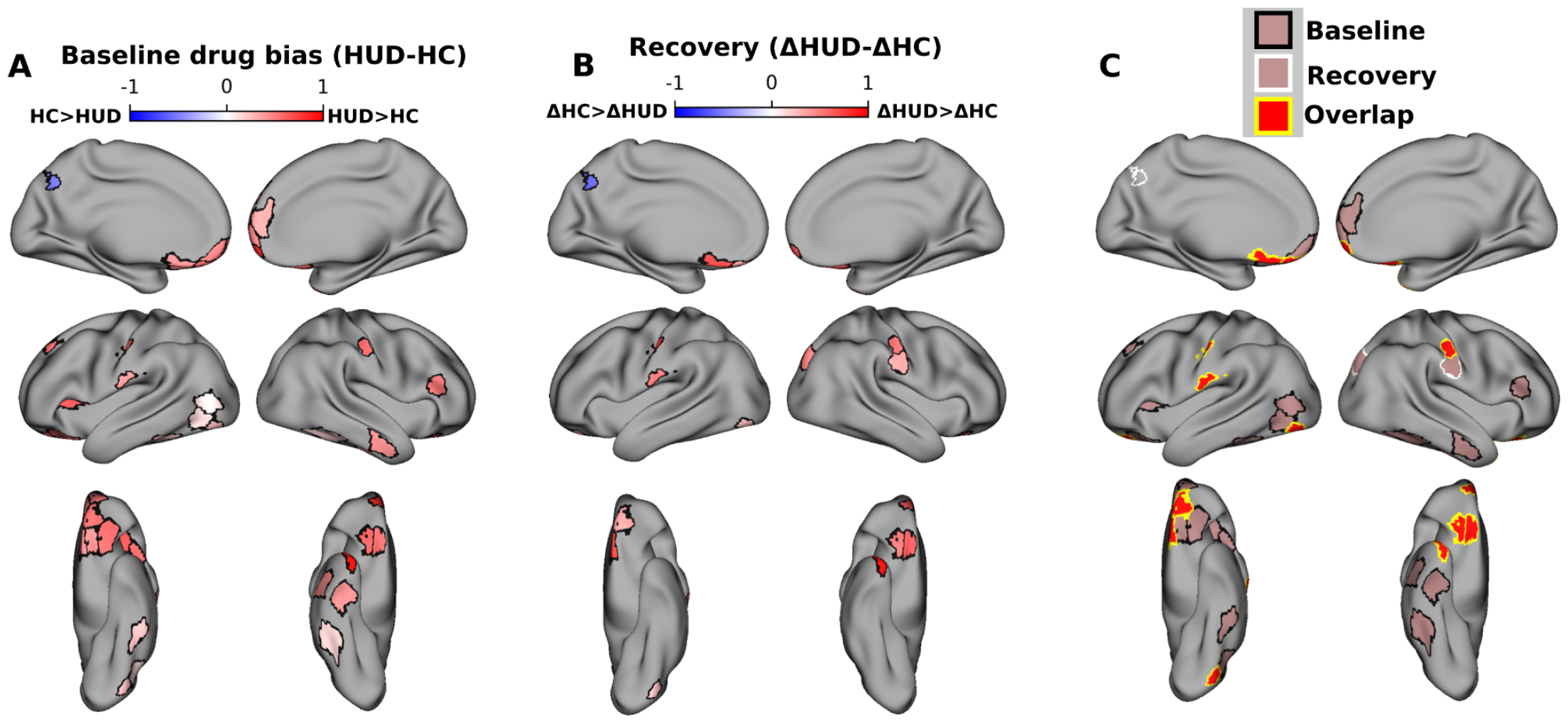
Drug-biased ROIs and recovery with treatment. A) ROIs with significant group differences in drug bias at baseline (FDR corrected at p<0.05). Colormap indicates the difference between groups (HUD minus HC) in the fraction of scenes preceding synchronized TRs that contained drug content. Red (blue) colors indicate significantly greater drug bias in HUD (HC). B) ROIs with significant group differences in changes in drug bias between sessions. Colormap indicates the difference between groups (HUD minus HC) of the difference between sessions (baseline minus follow up) in the fraction of scenes preceding synchronized TRs that contained drug content. Red (blue) colors indicate a significantly greater reduction in drug bias between sessions in HUD (HC). C) ROIs with significantly greater drug bias in HUD at baseline (black outline), significantly greater reduction in drug bias in HUD at follow up (white outline), and ROIs that showed both effects (yellow outline). Note that baseline and recovery tests were run independently whole brain and FDR-corrected over 450 ROIs.

### Drug-biased responses in HUD at baseline and reduction with treatment

At the baseline visit, reverse correlation analyses revealed significantly more drug bias in HUD compared to HC across 28 cortical ROIs, including several regions previously implicated in drug-cue reactivity and salience attribution (but also inhibitory control and higher-order executive function), such as the OFC, insula, and ventromedial, dorsolateral, and ventrolateral prefrontal cortex (Figure 1A); less drug bias in the HUD than HC group was observed in a single ROI (left posterior cingulate cortex, part of the default mode network).

To test for changes in drug bias between baseline and follow-up, within each group the baseline drug bias values were subtracted from their values at follow-up, repeating the permutation test for group differences in this delta. This test was again performed whole-brain over all 450 ROIs with FDR correction. Twelve ROIs, including several in the medial and lateral OFC, showed a significant group difference in the drug bias delta, whereby compared to HC, HUD exhibited a significant reduction from baseline to follow-up in this drug bias. Notably, ten ROIs, including five in the OFC, showed both a significantly greater drug bias at the baseline session and significantly more reduction with time in this bias in HUD compared to HC. Repeating these reverse correlation analyses using control labels for high brightness or loudness (i.e., testing for brightness or loudness bias) did not show any overlapping ROIs with the drug bias results (Fig S1), suggesting that these results are not driven generically by salient features of the stimulus. Similarly, control analyses suggest that these neural drug bias effects are not explained by group differences in sample characteristics (Table 1, see Supplement for the associated analyses).

### OFC inter-subject correlation (ISC) signal correlates with scene-induced craving

Scene-induced craving ratings in response to 3 second clips of the movie (sampled every 30 seconds, see Scene-specific post-movie survey in Methods) were obtained outside of the scanner and after the movie watching.. Importantly, in addition to the above described reduction in drug-biased brain responses between sessions, we observed a parallel significant reduction in this self-reported scene-induced craving in HUD (follow-up minus baseline session, paired test: -1.5±2.83, Wilcoxon rank sum=34.5, p=0.008, Table S1). We also collected craving ratings in the scanner immediately before and after the movie to measure craving induced by the movie as a whole; this more general movie-induced craving did not change significantly with treatment (Table S1).

Given the evidence for the role of the OFC in addiction and craving^3,21^, we next focused on the OFC to test whether changes in synchronized brain responses tracked changes in scene-induced craving ratings. For each session and for each ROI in the OFC, we extracted the ISC between each subject’s signal and the mean signal of the rest of their group^22^. We then averaged this ISC score over all OFC ROIs to obtain a single ISC score per subject-session. Finally, we tested correlations between these OFC-ISC scores and craving at the first session, as well as between the changes in both variables over time. There was a significant correlation between the change in OFC-ISC and the change in scene-induced craving (Pearson r: 0.54, p=0.0023, q=0.016 FDR corrected); at baseline there was a significant correlation that did not survive multiple comparison correction (Pearson r=0.38, p=0.04, q=0.14 FDR corrected; figure 3). We also tested correlations between OFC-ISC and several control measures, including the movie-induced craving, other measures obtained from the same post-movie survey that tested for the scene-induced craving, psychometric or demographic variables that differed between the groups, and measures related to heroin use other than craving (see correlation between OFC signal and control behavioral variables in supplement). These control measures did not change significantly between sessions (Table S2) and did not exhibit any significant correlations with the OFC-ISC scores (Table S6), suggesting a specific relationship between OFC synchronization and scene-induced craving.

**Figure 3.**
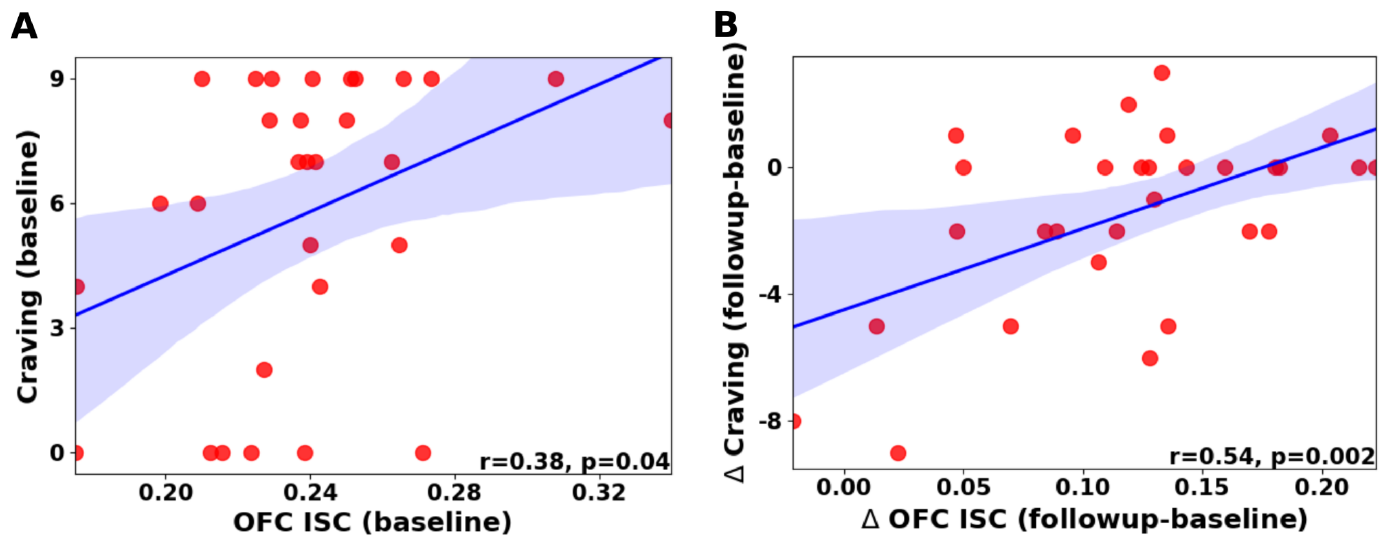
OFC ISC correlates with craving at baseline (A) and its delta tracks parallel changes in craving between sessions (B). Shaded area indicates 95% confidence interval for regression line.

## Discussion

For the first time using a naturalistic stimulus, where both drug and non-drug cues occupy a common narrative context, we found shared drug-biased reactivity in the OFC (and the vmPFC, vlPFC and insula, other regions within the reward, salience and inhibitory control networks) in iHUD, expanding on the limited evidence of such drug-biased processing in this population^3,23^. Further, for the first time we report reduction of this OFC cue reactivity with treatment-based abstinence in iHUD, indicative of recovery. Finally, this synchronized OFC response across subjects, and its treatment-induced changes, tracked respective changes in a dynamically derived drug craving, a more fine-tuned version of a well-established clinical outcome measure in addiction^16^. Our results therefore provide evidence for synchronized OFC responses as a neural marker of drug bias and craving, and their recovery, in HUD. Taken together, these results are consistent with previous drug cue reactivity studies across SUDs and with neurobiological theories of drug addiction that highlight OFC dysfunction in maladaptive reward processing, salience attribution and decision making^3,21,23,24^ such as the impaired response inhibition and salience attribution (iRISA) model^3^, extending them both clinically and into naturalistic neuroscience.

A classic theory of OFC function is that this region represents the value of expected outcomes of actions, particularly with respect to current metabolic and emotional needs^24^. More recent theories have reframed OFC function as contributing to the use of cognitive maps, which chart associations between metabolically and emotionally relevant states of the internal/external environment, in the way that physical maps chart associations between spatial locations^25,26^. A common thread across these theories is that the OFC supports model-based behavior, which privileges fast and flexible updates to behavior in novel environments^27^. Insofar as drug treatment strives to reduce the value of drug-related outcomes with respect to an individual’s current goals and needs, it is expected that the OFC would update (i.e., reduce) responses to drug cues with treatment^28^. Consistent with this role, theoretical accounts propose that OFC outcome value representations elicit feelings of craving^29^, which are supported by human imaging studies correlating OFC signals with craving ratings^21^, and more causal evidence for the role of the OFC in animal models of craving in addiction^30^. In line with these accounts of OFC function, we found that drug-biased OFC responses were reduced with treatment, whereby overall OFC synchronization tracked concomitant reductions in craving. While our results are promising vis-a-vis recovery of brain function with treatment, the long term efficacy of abstinence-based treatments is generally low^31^, suggesting that these OFC-based changes alone may be insufficient for maintaining long-term abstinence. Because OFC output is thought to be a primary learning signal for more habitual systems, e.g., in the striatum^32^, a major challenge in sustained recovery may relate to an individual’s ability to use an updated OFC cognitive map (or expected/model-based value signals) to retrain these model-free systems, which are inherently slower to learn.

Beyond exporting typical cue reactivity to a more naturalistic setting (allowing stimuli to compete for salience), our analyses take a fundamentally different view of what constitutes a response to drug content^22^. In particular, our analyses take into account the dynamics of brain responses, and show that during drug content these dynamics synchronize (show a shared response) across iHUD. While this complimentary view on drug cue reactivity largely aligns with previous picture-based studies in terms of the regions displaying group differences, there are also several regions, particularly the striatum, that were not detected by our method. However, we note that the level of synchronization in the striatum was low overall, suggesting either a high degree of idiosyncrasy across subjects or low signal quality in this region. Previous naturalistic studies have generally not analyzed subcortical areas, making it difficult to interpret our lack of subcortical findings without further experiments.

Several previous studies have focused on shared responses to naturalistic stimuli in other psychiatric conditions, predominantly through the lens of impaired social cognition in autism, major depressive disorder, or schizophrenia^33–41^. Our results can similarly be viewed as reflecting a social component of iRISA in that we specifically detect impairments in the collective group processing of drug cues. Speculatively, these shared impairments may facilitate collective behavior among peers at similar phases of the addiction cycle or recovery. Unlike these previous studies, however, our cue-reactivity analysis was based on a reverse correlation approach, which does not measure the overall level of synchronization between subjects, but rather allows inspection of the specific timing of synchronized responses to drug cues. Ours is also the first study to use this approach in a psychiatric condition in a longitudinal design (as a function of time into treatment). Showing the sensitivity of these synchronized brain patterns to treatment and relevant behavioral outcomes (here craving) further supports their use in psychiatry. Notably, as compared to a standard more static measure of craving assessed before and after the movie, the dynamic specific scene-induced craving measure was both more sensitive to treatment and brain synchronization (showing reduction with the former and correlation with the latter), suggesting that future studies could benefit from similar dynamic and more naturalistic sampling of behavior. In a related vein, an important novelty in our study is that the stimulus narrative was strongly related to addiction itself, a fine-tuning yet to be applied in naturalistic studies of other psychiatric conditions. An interesting avenue for future work is to compare responses to movies with clinically relevant narratives to those without and to identify batteries of movie stimuli that best probe the neural processes most relevant for a certain condition/disorder or even develop new naturalistic content explicitly for this purpose^8^.

Given the novelty of our approach in addiction neuroimaging, there remain several open questions. As we used a selected segment of a single movie, it will be important to test whether results generalize to other heroin-related movies in HUD. The use of additional movies would be crucial for solving three other potential confounds: (1) test-retest replication: we chose to repeat the same movie before and after an average of 15 weeks of treatment in the iHUD, while using the HC group to control for habituation (and other time-dependent) effects of repeated viewing. While previous studies have shown good test-retest replication of shared responses to naturalistic stimuli^7,42,43^, using different movies at both sessions could have addressed more directly the question of the possible impact on our results of viewing the same movie twice; (2) labeling of scenes (e.g., drug vs. non drug) in a complex movie: In picture-based cue reactivity studies, careful selection and matching of stimuli enhance the likelihood that only the dimension of interest (e.g., drug/non-drug cues) varies between different categories of pictures. In contrast, inherent in the use of rich naturalistic stimuli, such as an engaging movie where contents were not designed for our purposes a priori, is the possibility for characterizing any given moment or scene in a number of potentially salient or meaningful ways, where labels of interest can overlap, e.g., faces or social content and drug use (since people are using drugs together). Although we partially addressed this issue through testing alternative labels of the movie (see Figure S1), given this overlap, we cannot completely rule out the contribution of such features to the synchronization effects that we documented here, requiring the testing of additional movies where these features do not overlap. (3) statistical comparison with alternative reinforcers: while the movie used in this study was excellent for depicting drug use, it did not have enough scenes of alternative reinforcers or other arousing stimuli (food, sex, social) to run a direct statistical comparison between drugs and these categories. Similarly, arousal was not measured during movie watching and therefore we cannot rule out the impact of general arousal on our results.

Nevertheless, we took the important processing step of regressing out the global component from each voxel, which has been linked to general engagement and arousal to a movie stimulus^12^. While this step does not definitively remove the effects of arousal, it does strongly suggest that our results cannot be solely attributed to arousal.

Several other issues should be addressed with different population samples and pre-registered studies in the future. It will be important for future work to test a similar approach in other SUDs. Further, inferences about sex- and treatment-seeking status-related contributions warrant a larger sample size with more women and a non-treatment seeking group. Relatedly, our study does not allow for the decoupling between participation in the medication-assisted treatment (in addition to a randomized group therapy as part of the clinical trial, NCT04112186, see “Treatment-specific details” in supplement) and length of abstinence. While available self-reported methadone dosage did not change with treatment or correlate with outcomes of interest (Table S6), we cannot rule out the potential contribution of medication type (e.g., methadone vs. buprenorphine) or other treatment related measures. We also cannot test the contribution of group therapy type as part of the treatment received, to be evaluated separately at the completion of the clinical trial. However, the weekly group therapy effects are expected to be minor relative to the overall effect of inpatient treatment and differences between iHUD and HC. Finally, while the variables showing group differences (Beck’s Depression Inventory, Beck’s Anxiety Inventory, Fagerström Test for Nicotine Dependence, Short Michigan Alcohol Screening Test, years of regular marijuana use, verbal IQ, years of education, Table 1) did not correlate with outcomes of interest and thus do not explain the current results, samples more closely matched in demographics and smoking status are needed.

Taken together, using a naturalistic design in inpatients with HUD, we provide strong support for prominent theories of addiction^1,2^, including those that highlight the role of the OFC in salience attribution and craving^21^, paving a path towards identifying a dynamic, ecologically valid shared neural marker of recovery in this population. Future work validating these markers with other movies and groups with different SUDs will further help advance the field in this important direction.

## Methods

### Participants

Thirty iHUD (mean age=40.00±8.41 years; 6 women) were recruited from a medication assisted inpatient rehabilitation facility and 25 age- and sex-matched HC (mean age=39.97±10.76 years; 9 women) were recruited from the surrounding community for matching purposes through advertisements and word of mouth. See Table 1 for sample descriptive statistics and the supplement for exclusion criteria. The Icahn School of Medicine at Mount Sinai’s institutional review board approved study procedures, and all participants provided written informed consent. All iHUD were inpatients in a facility where they attended courses/treatments including relapse prevention, Seeking Safety therapy (a present-focused counseling model to help people attain safety from trauma and/or substance abuse), and anger management. A comprehensive clinical diagnostic and substance use interview was conducted at baseline, consisting of the Mini International Neuropsychiatric Interview 7th edition^44^ and the Addiction Severity Index 5th edition^45^. Drug dependence severity, craving, and withdrawal symptoms were assessed using the Severity of Dependence Scale^46^, the Heroin Craving Questionnaire (a modified version of the Cocaine Craving Questionnaire^47^) and the Short Opiate Withdrawal Scale^48^, respectively. Nicotine and alcohol dependence severity were measured using the Fagerström Test for Nicotine Dependence^49^ and the Short Michigan Alcohol Screening Test^50^, respectively. All iHUD met DSM-5 criteria for opioid use disorder (with heroin as the primary drug of choice/reason for treatment). Substance use-related psychiatric comorbidities commonly observed in individuals with drug addiction^51,52^ were either in partial or sustained remission at baseline with no current comorbidities found in the HC (see supplement). All iHUD were abstinent (182.90 ± 199.78 days during their baseline session) and under medication assisted treatment (confirmed via urine toxicology at both sessions and dosage in mg collected via self-report at baseline). Beck’s Depression and Anxiety Inventories and number of days since last heroin use were collected at both sessions. Table 1 details heroin route of administration, nicotine/alcohol/cannabis use, urine toxicology, and medication dose, in addition to demographics, neuropsychological and additional drug use measures. As part of the clinical trial associated with this study (NCT04112186), following baseline assessments including MRI, iHUD were randomly assigned to one of two types of group therapy (Mindfulness Oriented Recovery Enhancement or support group, both consisting of eight weekly two-hour sessions led by trained therapists. Therapy-specific results will be reported separately upon completion and unblinding of this ongoing clinical trial; see supplement for therapy group details). Data from participants in both treatment groups were combined for the present analyses to assess the general inpatient medication assisted treatment effect. Differences between the group therapies are expected to be minor relative to inpatient treatment overall and differences between HUD and HC, supporting the choice to combine treatment groups for the purposes of the current analyses.

### MRI acquisition and movie

MRI scans were acquired with a Siemens 3T Skyra (Siemens, Erlangen, Germany) and a 32-channel head coil while participants passively viewed the first 17 min 3 sec of the movie “Trainspotting.” The video was played on a television outside the MRI bore, which participants viewed via a mirror mounted on the head coil, while MRI-compatible in-ear headphones were used for the audio. The MRI protocol was optimized to be Human Connectome Project compatible^53^. BOLD fMRI responses were assessed via a T2*-weighted single-shot multi-band (acceleration factor of 7) gradient-echo EPI sequence (TE/TR=35/1000 ms), with 2.1 mm isotropic resolution, 70 axial slices for whole brain coverage (14.7 cm), 206 × 181 mm FOV, 96 × 84 matrix size, 60°-flip angle, blipped CAIPIRINHA phase-encoding shift=FOV/3, ∼2 kHz/pixel bandwidth with ramp sampling, 0.68 ms echo spacing, and 57.1 ms echo train length. T1-weighted anatomical scans were acquired using a 3D MPRAGE sequence (TR/TE/TI=2400/2.07/1000 ms) with 0.8 mm isotropic resolution, 256 × 256 × 179 mm^3^ FOV, 8° flip angle with binomial (1, −1) fat saturation, 240 Hz/pixel bandwidth, 7.6 ms echo spacing, and an in-plane acceleration (GRAPPA) factor of 2. The scan session included other procedures unrelated to the movie reported elsewhere^11,23,54–57^. Immediately before and after the movie, subjects were asked to rate their desire for heroin on a scale of 0-9.

### Scene-specific post-movie survey

Outside of the scanner (within 45 minutes after watching the movie), each subject completed scene-specific craving ratings. For this purpose, we extracted 3-sec clips separated by regular 30 sec intervals to densely sample movie scenes in an unbiased manner. This yielded 34 3-sec clips in the 17-min movie. Participants were instructed to watch each clip and provide subjective ratings of their experience when they watched that clip in the scanner, including their craving, and any scene-induced emotion amplification or suppression (for purposes of measuring savoring vs. reappraising, respectively, not related to the current goals). For correlations with brain data, the maximum of these scene-by-scene ratings was taken for each subject, a measure that accounts for natural variation in the stimulus features that elicit craving. The survey also included several questions to ascertain comprehension and memory including whether the participant had previously watched the movie, self-report ratings of attention during movie-watching, understanding of the movie, ratings of perceived audio and video quality, followed by a four-question memory probe related to the general theme of the movie: “On which drug is the movie primarily centered?”, “Is the main character trying to quit using a drug?”, “Did the main character and his friends live in rich neighborhoods?”, and “Did anyone in the movie die of drug-related causes, and if so, how many?”. The survey also included a recognition task comprising a series of drug and non-drug word stimuli representing target and distractor items, where participants were asked whether they had seen each item in the movie, and how confident they felt about their response. There were no significant group differences in these comprehension and memory questions.

### Anatomical data preprocessing with fMRIprep version 20.2.1^58^

For each subject, a T1-weighted (T1w) image was corrected for intensity non-uniformity with N4BiasFieldCorrection^59^, distributed with ANTs 2.3.3^60^ and used as T1w-reference throughout the workflow. The T1w-reference was then skull-stripped with a Nipype implementation of the antsBrainExtraction.sh workflow (from ANTs), using OASIS30ANTs as the target template. Brain tissue segmentation of cerebrospinal fluid, white-matter and gray-matter was performed on the brain-extracted T1w using FAST [FSL 5.0.9, RRID:SCR_002823^61^]. Volume-based spatial normalization to a standard space (MNI152NLin2009cAsym) was performed through nonlinear registration with antsRegistration (ANTs 2.3.3), using brain-extracted versions of both T1w reference and the T1w template. The ICBM 152 Nonlinear Asymmetrical template version 2009c^62^ [RRID:SCR_008796; TemplateFlow ID: MNI152NLin2009cAsym] was used for spatial normalization.

### Functional data preprocessing with fMRIPrep version 20.2.1

For all BOLD movie runs (across all subjects), the following preprocessing was performed. First, a reference volume and its skull-stripped version were generated by aligning and averaging a single-band reference. A B0-nonuniformity map (or fieldmap) was estimated based on two echo-planar imaging (EPI) references with opposing phase-encoding directions, with AFNI’s 3dQwarp^63^. Based on the estimated susceptibility distortion, a corrected EPI (echo-planar imaging) reference was calculated for a more accurate co-registration with the anatomical reference. The BOLD reference was then co-registered to the T1w reference using FLIRT [FSL 5.0.9^64^] with the boundary-based registration^65^ cost-function. Co-registration was configured with nine degrees of freedom to account for distortions remaining in the BOLD reference. Head-motion parameters with respect to the BOLD reference (transformation matrices, and six corresponding rotation and translation parameters) were estimated before any spatiotemporal filtering using MCFLIRT [FSL 5.0.9^66^]. First, a reference volume and its skull-stripped version were generated using a custom methodology of fMRIPrep. The BOLD time-series were resampled onto their original, native space by applying a single, composite transform to correct for head-motion and susceptibility distortions. The BOLD time-series were resampled into the MNI152NLin2009cAsym standard space and used for further custom preprocessing.

### Custom preprocessing

Spatially-normalized, preprocessed BOLD data in the MNI152NLin2009cAsym standard space were further processed with the following steps. A gray-matter mask was generated by averaging tissue probability maps of all subjects, then binarizing the resulting map by thresholding at 95% probability gray-matter. The gray-matter mask and gaussian smoothing (6 mm) were then applied. In the remaining gray-matter voxels, the first 10 TR’s were removed from all BOLD time series to eliminate contributions of large stimulus onset responses to further analyses^22^. The following confounds were regressed out of each BOLD signal: 6 translation and rotation parameters (x,y,z for each), their square, their derivative, and their squared derivative, as well as the cerebrospinal fluid component output by fMRIPrep. Regression of confounds, high-pass filtering (period of 140 s), linear detrending, and z-scoring were applied in a single step using the signal.clean function from nilearn Python package^67^.

### Global component and selective components

For each subject the preprocessed BOLD time series were averaged over all gray-matter voxels and then z-scored, resulting in a single global component per subject. Selective components were then derived by regressing the time series at each voxel within a subject onto the global component for that subject with a least-squares linear model. The residual of this linear fit was then z-scored and kept as the selective component for that subject-voxel^12^.

### Parcellations and shared response model

Cortical ROIs were selected by applying the Schaefer functional parcellation^68^ with 400 regions. Subcortical ROIs were selected with the Melbourne Subcortical Atlas^69^ with 50 regions. To simplify analyses, and focus on shared group responses, we derived a single time series per ROI for each subject by applying a shared response model with a single shared component per group^70^. This method projects the data for each subject to a single component that maximally explains the observed voxel-wise data within each ROI via a linear transform. The shared response model was implemented via the BrainIAK toolbox^71^. All further analyses were conducted on this shared component within each ROI.

### Identifying synchronized TR’s and region-specific reactivity

Within each ROI and group we aimed to identify individual TRs exhibiting significant synchronization across individuals, indicated by peaks in the group median of the shared component. At each TR we estimated 95% confidence intervals for the group median via 5000 subject-wise bootstraps. We then compared this confidence interval for the median to the maximum value that could be expected by chance synchronization. We performed 5000 phase randomizations of the shared component (by taking the FFT, randomizing the phase of all frequency components, and then taking the inverse FFT), calculated the median signal and kept the maximum value over all TRs from each randomization to build a null distribution. A TR had a significant response if the 95% confidence interval for the group median was above the 95th percentile of the null distribution. This yielded a set of significant TRs, T*={t_i_}, which reflects synchronized activity within a given group of subjects in a specific ROI.

To gain insight into the movie content that drove the synchronization in a group of subjects, T*, we performed a reverse correlation-like analysis (inspired by analysis of single unit recordings) similar to Hasson et al.^12^ For each t_i_ in T*, we collected a 5 second clip of the movie corresponding to t_i_-10 to t_i_-5 to account for the delay in the hemodynamic response. Each 5 second clip was split into 1 second bins that were labeled drug/non-drug (see movie labeling). This procedure mapped T* to the fraction of unique time bins that were labeled as drug (out of total bins), which we called the drug bias and used as a test statistic. For contrasts between groups, p-values were obtained by taking the difference between the drug bias for each group and comparing this difference to a null distribution generated by randomizing T* for each group 5000 times. For group-by-session interaction effects, the test statistic was the difference in drug bias between sessions, which was then compared between groups. The same procedure was repeated for brightness and loudness labels (see movie labeling).

While our approach here is inspired by that of Hasson et al.^12^, and is conceptually very similar, we made several changes primarily to reduce the rate of false positives in identifying synchronized TRs. In particular, the original paper of Hasson et al. ran independent t-tests at each TR without correcting for multiple comparisons over the number of TRs.

Here we took a nonparametric approach to avoid assumptions about the sample distribution and controlled the family-wise error rate by comparing to a null distribution of maximum values over 5000 phase randomizations. Our method of statistically comparing the movie labels that resulted from the reverse correlation is also novel, as Hasson et al.’s assessment of the resulting movie frames was qualitative.

### Movie labeling

To map synchronized TRs to movie content, we first binned the movie into 1 second clips with the start and end of each clip aligned to a TR. Each 1 second clip was then labeled by raters blind to the current analyses/results as drug if it contained any drugs, drug use, or a character who is high; and labeled as non-drug otherwise. This yielded 464 seconds labeled drug out of 1023 seconds total (45.4%). As a control we also created loudness and brightness labels, which were extracted using the pliers package^72^. The route-mean-square of the audio signal and the average luminosity of the pixels was averaged within each 1 second bin. Bins were then labeled as high or low loudness/brightness by a median split over all bins.

### Correlations with behavior

Inter-subject correlation in the OFC was used as a subject level brain signal for all correlations with behavior. For these analyses, we considered all ROIs labeled as OFC by the Schaefer parcellation^68^. Within each OFC ROI, ISC was calculated per subject taking the Pearson correlation between their shared response and the mean response of the rest of the group for that session. The ISC scores were then Fisher z transformed, averaged over all OFC ROIs, and inverse transformed to get a single OFC ISC score per subject. These ISC scores, or their difference between sessions, were then correlated with four different craving measures: maximum scene-induced craving, pre-movie craving, movie-induced craving, and the Heroin Craving Questionnaire total score (see Supplement for details). Statistical significance was determined after applying FDR correction over all correlations between OFC ISC and craving measures. We further tested correlations between OFC ISC and several other control measures that either contained behavioral responses to the movie, differed between groups, or were related to drug use, but unrelated to craving (see Supplement for details).

## Data Availability

All data produced in the present study are available upon reasonable request to the authors.

## Acknowledgements

We thank Uri Hasson for his helpful comments and discussions of this manuscript.

## Supplement

### Exclusion criteria

Exclusion criteria for all participants were the following: 1) DSM-5 diagnosis for schizophrenia or neurodevelopmental disorder; 2) Head trauma with loss of consciousness longer than 30 min; 3) History of central nervous system disease; 4) Cardiovascular, metabolic, endocrinological, oncological, autoimmune, and infectious diseases including Hepatitis B and C or HIV/AIDS; 5) Metal implants or other MR contraindications (including pregnancy); 6) Court mandated treatment. We did not exclude for DSM-5 diagnosis of a drug use disorder other than opiates as long as heroin was the primary drug of choice/reason for treatment-seeking since iHUD commonly use alcohol, amphetamines, benzodiazepines, other sedatives, cocaine, and marijuana in addition to heroin. Exclusion criteria for the HC were the same, except history of any drug use disorder was prohibitive.

### Diagnostic details in individuals with heroin use disorder

Comorbidities in iHUD included major depressive disorder (n=6), cocaine use disorder (n=5), post-traumatic stress disorder (n=2), sedative use disorder (n=3), cannabis use disorder (n=2), alcohol use disorder (n=3), generalized anxiety disorder (n=1), meth/amphetamine use disorder (n=1), and panic disorder (n=1).

### Controlling for group differences in sample characteristics

The HUD and HC groups differed significantly in years of education, verbal IQ, Beck’s Depression Inventory, Beck’s Anxiety Inventory, Fagerström Test for Nicotine Dependence, the Short Michigan Alcohol Screening Test and years of regular marijuana use (Table 1). To inspect their putative contribution to results, we tested these variables’ correlations with the root-mean-square signal at significant time points (see Identifying synchronized TR’s and region-specific reactivity) in the ROIs that showed higher drug bias in HUD at baseline (yielding 392 tests: 28 ROIs x 7 control variables x 2 groups). None of these ROIs showed a significant correlation when correcting for multiple comparisons (with FDR correction). Similarly, we tested correlations between each of these demographic variables and ISC scores in the OFC and did not find significant correlations (Table S6). Overall, these control analyses suggest that our main results (the drug bias measured here and the OFC as a marker of craving) were not driven by group differences in the above variables.

### Treatment-specific details

Study participants were randomized into Mindfulness Oriented Recovery Enhancement and support group therapy sessions, both received in addition to medication-assisted inpatient treatment. The former group sessions involved mindfulness-based self-awareness and emotion regulation training (e.g., cognitive reappraisal of negative and savoring of positive thoughts/contexts). These strategies were delivered with the goal of diminishing drug cue-reactivity while enhancing natural reward processing and cognitive control over craving and drug-seeking behaviors (see^73^ for therapy details). The support group included structured, therapist-guided and addiction-related psychoeducation, emotional expression, and discussions. In both groups, subjects were instructed to supplement the group sessions with daily 15-min independent practice sessions guided by audio instructions or journaling, respectively.

### Reverse correlation with control movie labels

To test for generic effects of salient movie labels on reverse correlation results, we generated two sets of control labels for the movie, one for high/low brightness and one for high/low loudness (see methods for labeling details). Repeating the whole-brain reverse correlation analyses with these labels revealed group differences between HUD and HC, but none overlapped with the ROIs identified by the drug bias analysis. This result suggests that the drug bias effects cannot be explained by labeling any salient feature of the movie.

**Supplemental Figure 1.**
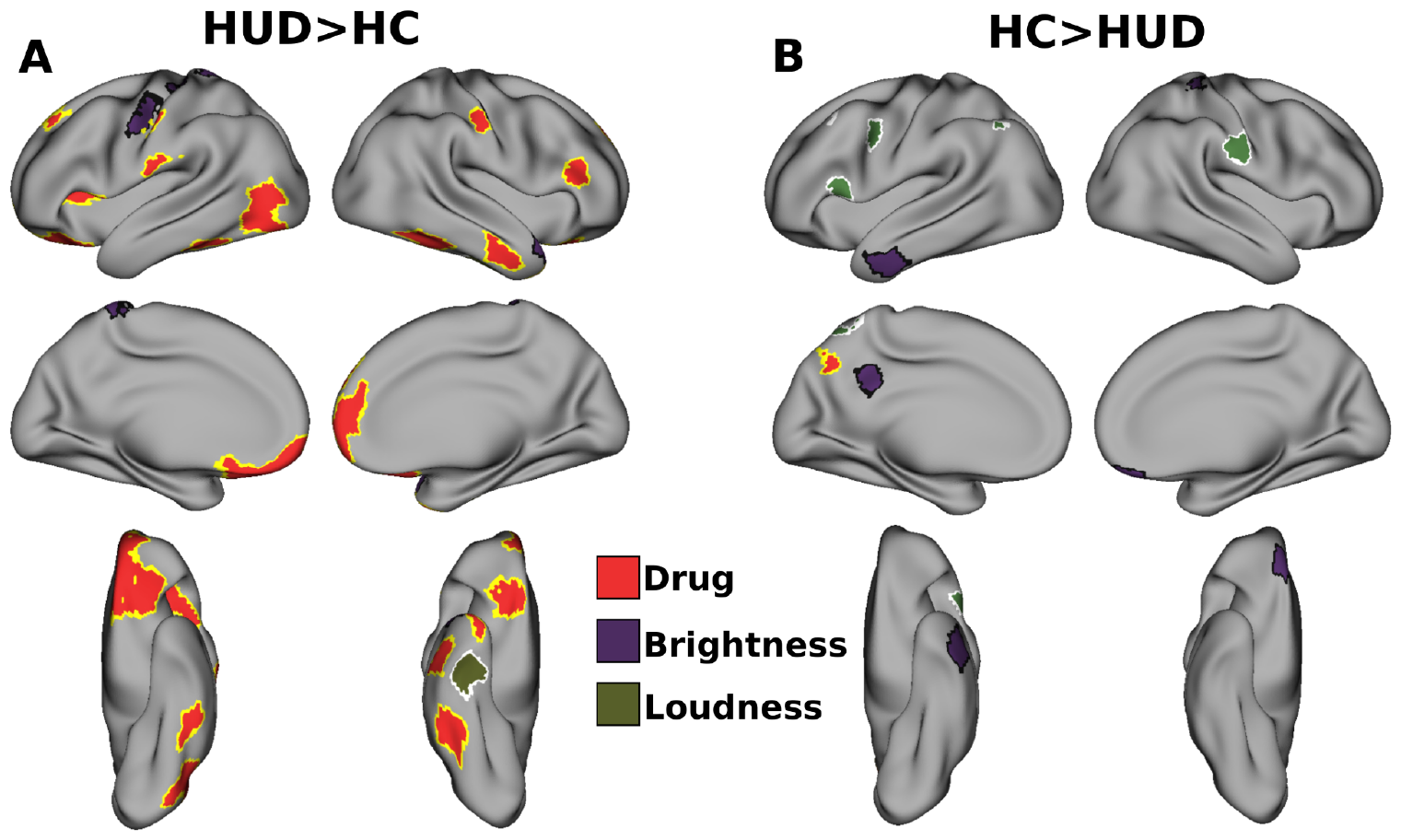
Reverse correlation maps with loudness and brightness labels do not overlap with drug labels. ROIs showing significantly higher bias in HUD (A) or HC (B) towards drug (red), high brightness (purple), or high loudness (green) content at the baseline session. Note that significant ROIs do not overlap between labels, particularly in the OFC.

### Testing changes in control measures between sessions

Our primary behavioral outcome of interest was craving. We had three craving measures collected both before and after treatment: scene-induced craving, pre-movie craving, and movie-induced craving; a fourth measure, the Heroin Craving Questionnaire, was only collected at baseline. Given that we only expect meaningful craving signals in HUD (no HC participants reported drug craving in any of these measures), we performed one-sample paired tests (follow-up minus baseline) in HUD for each of these craving measures; the resulting p-values were corrected for multiple comparisons with FDR correction. Only scene-induced craving showed a significant reduction after treatment (Table S1). Although the movie was drug-related, this result suggests that fine-grained scene-level craving, which more closely assesses cue-reactive dynamic fluctuations in craving, are needed to demonstrate changes in craving with treatment; these same craving measures were also the only to correlate with the OFC (main text). To test the specificity of craving in iHUD as a behavioral marker of treatment outcome, we also ran one-sample paired tests on five other control measures that were collected at both sessions and found no other significant changes with treatment in HUD (Table S2) or in HC (Table S3, S4).

**Table S1.** Statistical tests for between session craving effects in HUD. Paired (follow-up minus baseline) tests were performed against the null hypothesis of zero mean. q indicates FDR-corrected p-values.

| Craving variable | test | statistic | p | mean | std | q |
| --- | --- | --- | --- | --- | --- | --- |
| Scene-induced craving | wilcoxon | 34 | 0.008 | -1.5 | 2.8 | 0.024 |
| Pre-movie craving | ttest | -2 | 0.052 | -0.74 | 1.9 | 0.072 |
| Movie-induced craving | wilcoxon | 29 | 0.072 | -0.56 | 1.5 | 0.072 |

**Table S2.** Statistical tests for between session effects in control variables in HUD. Paired (follow-up - baseline) tests were performed against the null hypothesis of zero mean. q indicates FDR-corrected p-values.

| Control variable | test | statistic | p | mean | std | q |
| --- | --- | --- | --- | --- | --- | --- |
| Scene-induced emotional increase | ttest | -0.92 | 0.37 | -0.63 | 3.7 | 0.62 |
| Scene-induced emotional decrease | ttest | -0.22 | 0.83 | -0.13 | 3.2 | 0.83 |
| Movie item recognition | ttest | -1.5 | 0.14 | -0.033 | 0.12 | 0.35 |
| Beck's Depression Inventory | ttest | -1.8 | 0.09 | -2.8 | 8.3 | 0.35 |
| Beck's Anxiety Inventory | ttest | 0.48 | 0.63 | 0.69 | 7.6 | 0.79 |
| Methadone dose | ttest | 0.0 | 1.0 | 0.0 | 26.0 | 1.0 |

**Table S3.** Statistical tests for between session craving effects in HC. Paired (follow-up - baseline) tests were performed against the null hypothesis of zero mean. q indicates FDR-corrected p-values.

| Craving variable | test | statistic | p | mean | std | q |
| --- | --- | --- | --- | --- | --- | --- |
| Scene-induced craving | wilcoxon | 2.5 | 0.79 | -0.042 | 1.2 | 0.79 |
| Pre-movie craving | wilcoxon | 0 | 0.32 | -0.21 | 1 | 0.48 |
| Movie-induced craving | ttest | 1.7 | 0.1 | 0.83 | 2.4 | 0.3 |

**Table S4.** Statistical tests for between session effects in control variables in HC. Paired (follow-up - baseline) tests were performed against the null hypothesis of zero mean. q indicates FDR-corrected p-values.

| Control variable | test | statistic | p | mean | std | q |
| --- | --- | --- | --- | --- | --- | --- |
| Scene-induced emotional increase | ttest | -1.6 | 0.12 | -1.4 | 4 | 0.6 |
| Scene-induced emotional decrease | wilcoxon | 15 | 0.37 | 0.5 | 2.4 | 0.64 |
| Movie item recognition | ttest | 0.8 | 0.43 | 0.015 | 0.091 | 0.64 |
| Beck's Depression Inventory | ttest | 0.48 | 0.64 | 0.28 | 2.9 | 0.64 |
| Beck's Anxiety Inventory | ttest | 0.56 | 0.58 | 0.32 | 2.8 | 0.64 |

**Table S5.** Correlations between OFC ISC and craving measures in HUD. For rows involving deltas between sessions, the difference between follow-up and baseline was used for both the craving measures and OFC ISC scores. r indicates Pearson correlation coefficient, q indicates FDR-corrected p values.

| Craving variable | Session | r | p | q |
| --- | --- | --- | --- | --- |
| Scene-induced craving | Baseline | 0.38 | 0.04 | 0.14 |
| Pre-movie craving | Baseline | 0.29 | 0.12 | 0.27 |
| Movie-induced craving (post-pre) | Baseline | -0.15 | 0.44 | 0.61 |
| Heroin Craving Questionnaire | Baseline | 0.079 | 0.68 | 0.68 |
| Scene-induced craving | Delta (follow-up - baseline) | 0.54 | 0.0023 | 0.016 |
| Pre-movie craving | Delta (follow-up - baseline) | 0.12 | 0.54 | 0.63 |
| Movie-induced craving (post-pre) | Delta (follow-up - baseline) | -0.27 | 0.16 | 0.27 |

**Table S6.** Correlations between OFC ISC and control measures in HUD. For rows involving deltas between sessions, the difference between follow-up and baseline was used for both the craving measures and OFC ISC scores. r indicates Pearson correlation coefficient, q indicates FDR-corrected p values.

| Control variable | Session | r | p | q |
| --- | --- | --- | --- | --- |
| Scene-induced emotional increase | Baseline | 0.15 | 0.44 | 0.93 |
| Scene-induced emotional decrease | Baseline | -0.034 | 0.86 | 0.93 |
| Movie item recognition | Baseline | 0.028 | 0.88 | 0.93 |
| Beck's Depression Inventory | Baseline | 0.28 | 0.14 | 0.93 |
| Beck's Anxiety Inventory | Baseline | 0.046 | 0.81 | 0.93 |
| Fagerström Test for Nicotine Dependence | Baseline | -0.036 | 0.85 | 0.93 |
| Short Michigan Alcohol Screening Test | Baseline | -0.0096 | 0.96 | 0.96 |
| Years of regular marijuana use | Baseline | -0.22 | 0.26 | 0.93 |
| Verbal IQ | Baseline | 0.035 | 0.86 | 0.93 |
| Years of Education | Baseline | 0.22 | 0.26 | 0.93 |
| Years of regular heroin use | Baseline | -0.34 | 0.073 | 0.73 |
| Heroin Severity of Dependence Scale | Baseline | -0.13 | 0.5 | 0.93 |
| Short Opiate Withdrawal Scale | Baseline | 0.039 | 0.84 | 0.93 |
| Days of abstinence from heroin | Baseline | 0.054 | 0.78 | 0.93 |
| Days of heroin use in past 30 days | Baseline | 0.048 | 0.81 | 0.93 |
| Methadone Dose | Baseline | 0.33 | 0.13 | 0.73 |
| Scene-induced emotional increase | Delta (follow-up-baseline) | -0.12 | 0.54 | 0.93 |
| Scene-induced emotional decrease | Delta (follow-up-baseline) | -0.17 | 0.37 | 0.93 |
| Movie item recognition | Delta (follow-up-baseline) | -0.12 | 0.51 | 0.93 |
| Beck's Depression Inventory | Delta (follow-up-baseline) | 0.43 | 0.021 | 0.42 |
| Beck's Anxiety Inventory | Delta (follow-up-baseline) | 0.061 | 0.75 | 0.93 |
| Methadone dose | Delta (follow-up-baseline) | 0.43 | 0.087 | 0.62 |

### Correlation between OFC signal and control behavioral variables

We hypothesized that OFC ISC scores would specifically correlate with measures of craving. We had four different craving measures at baseline (scene-induced craving, pre-movie craving, movie-induced craving, and the Heroin Craving Questionnaire). Of these craving measures, scene-induced and pre-movie craving were intercorrelated at baseline (r:0.68, p:0.000045) as well as in their delta between sessions (r:0.6, p:0.00093). We tested for correlations between all four of these craving measures and OFC ISC scores at baseline and for correlations between the session deltas of each of these craving measures (except the Heroin Craving Questionnaire that was only collected at baseline) and the session deltas of the OFC ISC scores. FDR correction was applied over the seven correlations tested. Only the correlation between the session deltas with scene-induced craving survived multiple comparisons correction (q<0.05); at baseline, scene-induced craving showed a trend toward significance (p<0.05) (Table S5).

To test the specificity of the relationship between OFC ISC scores and craving, we also ran correlations between the OFC ISC scores and several control measures. Here we included three measures from the same post-movie survey used for the scene-induced craving measure (ratings for whether a subject was able to increase or decrease their emotional reaction to a scene, item recognition memory scores), demographic or psychometric variables that differed between the groups (Beck’s Depression Inventory, Beck’s Anxiety Inventory, Fagerström Test for Nicotine Dependence, Short Michigan Alcohol Screening Test, years of regular marijuana use, verbal IQ, years of education), and measures other than craving that specifically related to heroin use (years of regular heroin use, Heroin Severity of Dependence Scale, Short Opiate Withdrawal Scale, days of abstinence from heroin, days of heroin use in the past 30 days). We also tested correlations between the session deltas of OFC ISC scores and the control variables that were collected at both sessions. FDR correction was applied over all control correlations tested. None of these control variables survived FDR correction (q<0.05), although the session delta for Beck’s Depression Inventory showed a trend towards significance (p<0.05).

